# The effect of sleep-corrected social jetlag on crystalized intelligence, school performance, and functional connectome in early adolescence

**DOI:** 10.1101/2023.07.18.23292833

**Authors:** Fan Nils Yang, Dante Picchioni, Jeff H. Duyn

## Abstract

Approximately half of adolescents encounter a mismatch between their sleep patterns on school days and free days, also referred to as “social jetlag”. This condition has been linked to various adverse outcomes, such as poor sleep, cognitive deficits, and mental disorders. However, prior research was unsuccessful in accounting for other variables that are correlated with social jetlag, including sleep duration and quality. To address this limitation, we applied a propensity score matching method on a sample of 8853 11-12-year-olds from the two-year follow-up (FL2) data of the Adolescent Brain Cognitive Development (ABCD) study. We identified 3366 pairs of participants with high sleep-corrected social jetlag (SJLsc, over 1 hour) and low SJLsc (<= 1 hour) at FL2, as well as 1277 pairs at three-year follow-up (FL3), after matching based on 11 covariates including socioeconomic status, demographics, and sleep duration and quality. Our results showed that high SJLsc, as measured by the Munich Chronotype Questionnaire, was linked to reduced crystallized intelligence, lower school performance - grades, and decreased functional connectivity between cortical networks and subcortical regions, specifically between cingulo-opercular network and right hippocampus (cerc-hprh). Further mediation and longitudinal mediation analyses revealed that cerc-hprh connection mediated the associations between SJLsc and crystallized intelligence at FL2, and between SJLsc and grades at both FL2 and FL3. We validated these findings by replicating these results using objective SJLsc measurements obtained via Fitbit watches. Overall, our study highlights the negative association between social jetlag and crystallized intelligence during early adolescence.

## Introduction

In today’s fast-paced and interconnected world, the prevalence of social jetlag (SJL) has become a significant health concern. SJL refers to the misalignment between an individual’s biological clock and their social schedules (Roenneberg et al., 2003; Wittmann et al., 2006). While SJL presents throughout work life, adolescents often experience the most severe SJL (Roenneberg et al., 2019), as their biological clock progressively shifts to the late phase during adolescence (Fischer et al., 2017). SJL has been associated with mood disorders, aggression and behavioral conduct problems, cognitive problems, and metabolic diseases in both adults (Beauvalet et al., 2017; Caliandro et al., 2021; Roenneberg et al., 2019; Taillard et al., 2021) and adolescents (Chen et al., 2022; Díaz-Morales & Escribano, 2015; Henderson et al., 2019; Tamura et al., 2022). However, due to significant heterogeneity between study methodologies, the direct effects of SJL on behavior and brain are poorly understood. First, SJL significantly correlated with other sleep measurements, i.e., sleep duration and quality, as well as socioeconomic status. These factors typically have not been considered concurrently in previous studies. Given that our recent studies show that inadequate sleep is also related to mood disorders, crystallized intelligence, and impulsivity (Yang et al., 2023; Yang, Liu, et al., 2022; Yang, Xie, et al., 2022), it was impossible to measure direct effects of SJL on above-mentioned behaviors unless other sleep measures are carefully controlled. Second, the calculation of SJL is inconsistent across studies. Classic calculation of SJL failed to consider oversleeping during weekends, which is common among adolescents (Henderson et al., 2019; Tamura et al., 2022). To account for the latter, a sleep-corrected SJL (SJLsc) takes the weekday-weekend sleep duration difference into account (Jankowski, 2017). Third, most studies relied on self- or parent-reported sleep measures, which might overestimate child sleep as compared to objective sleep measures (Dayyat et al., 2011; Perpétuo et al., 2020), such as actigraphy. In addition, only a few studies investigated the functional connectivity associations of SJL in adults (Jia et al., 2023; Nechifor et al., 2020, 2022), and neural associations of SJL/SJLsc in adolescents are poorly understood.

To address these shortcomings, we applied propensity score matching (PSM) to investigate how SJLsc, measured by Munich Chronotype Questionnaire (MCTQ), is associated with functional connectivity and behavior in early adolescents after controlling for various covariates, such as other sleep measurements, i.e., sleep duration and quality, socioeconomic status, Body Mass Index (BMI), puberty status, etc. We then verified the results using an objective measurement of SJLsc through wearable devices. Based on previous research, we hypothesized that SJLsc would have adverse effects on behavioral problems, cognitive performance, mental health, and brain functional connectivity. We predicted that identified functional connectivity would mediate the association between SJLsc and behavioral measures.

## Methods

### Study design and data source

In this longitudinal, observational cohort study, we utilized data from a population-based sample of 9-10-year-olds (11-13 year-olds onward in the current study) collected from 21 study sites across the United States as part of the ongoing Adolescent Brain Cognitive Development (ABCD) study. The data used in our study were obtained from the ABCD data release 4.0, which encompassed baseline data collected between September 1, 2016, and October 15, 2018, as well as follow-up data collected from July 30, 2018, to January 15, 2021. This comprehensive dataset included behavioral and neural information gathered at baseline, 1-year, 2-year, and 3-year follow-ups. Only data from 2-year and 3-year follow-ups were used in the current study. The study employed rigorous protocols and stratified sampling techniques to ensure accurate data collection and representation of the diverse United States population.Detailed protocols and designs were described previously (Casey et al., 2018). Participants with missing data for relevant covariates used in PSM were excluded (see Appendix 1.1 for details). This resulted in 8853 children out of 10414 children. Informed consent was obtained from primary caregivers and assent from children. The study was approved by the Institutional Review Boards of all 21 study sites.

### Subjective sleep measures

Starting from the 2-year follow-up (FL2), Munich Chronotype Questionnaire (MCTQ) was administered (Roenneberg et al., 2003, 2019; Wang et al., 2023). Sleep duration (SDweek) and SJLsc were derived from this MCTQ data. SDweek reflects average sleep duration across the whole week, i.e., school days and free days. Classic SJL was calculated as the absolute value of the mid sleep (defined as midpoint between sleep onset (SO) and wake up) difference between work/school days (MSW) and weekends/free days (MSF) (equation 1). However, classic SJL did not take the potential sleep deprivation on school days and/or oversleeping on free days into account (Jankowski, 2017; Roenneberg et al., 2019). This effect can be minimized by SJLsc (Jankowski, 2017), which takes SD_week_ into account.

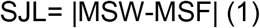

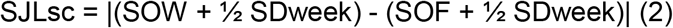

Here, in SJLsc calculation (equation 2), instead of using MSW/MSF it added half of the average sleep duration across the week to the sleep onset on both work/school day (SOW) and free days (SOF). It should be noted that SJLsc actually reflects the difference in sleep onset between school days and free days. For quality control purposes, we excluded children whose reported sleep duration of school days/free days were longer than 15 hours or shorter than 3 hours per day (299 out of 9153 eligible children), because these abnormal sleep duration were more likely caused by mistakes that happened in data collection processes. Total sleep disturbance score (TSD) is measured by the parent-reported Sleep Disturbance Scale for Children, which reflects the sum score of six sleep disorders, including disorders of initiating and maintaining sleep, sleep breathing disorders, disorder of arousal, sleep–wake transition disorders, disorders of excessive somnolence, and sleep hyperhidrosis.

### Objective sleep measures

Actigraphy data, i.e., Fitbit Charge HR, were collected and processed at FL2 by the Data Analysis, Informatics, and Resource Centre of the ABCD study. A subset of children (n = 5901) were instructed to wear Fitbit watches for 3 consecutive weeks after the onsite visit. We included data from 3821 children who had at least 7 nights (Aili et al., 2017; Giddens et al., 2022) of sleep recording within three weeks of the onsite visit and had at least 3 nights of sleep recording on school days/free days. Objective SJLsc (oSJLsc) was calculated based on data recorded from Fitbit watches. For quality control purposes, we excluded sleep duration data that were longer than 15 hours or shorter than 3 hours.

### Behavioral problems, cognition, mental health, and school performance measures

*Behavioral problems*: We used the parent-reported Child Behaviour Checklist at FL2 and 3-year follow-up (FL3) to measure a child’s behavioral problems in emotional, social, and behavioral domains; *Cognition*: we used scores from the US National Institutes of Health (NIH) Cognition Battery Toolbox at FL2 to assess a child’s cognitive functions. However, 4 out of 10 cognitive measures were not collected at FL2. *Mental Health*: We estimated a child’s overall mental health at FL2 based on scores from the brief child version of the Prodromal Psychosis Scale (PPS); the youth version of the Urgency, Premeditation (lack of), Perseverance (lack of), Sensation Seeking, Positive Urgency (UPPS-P) Impulsive Behaviour Scale; and the Behavioural Inhibition Scale (a summary of these variable names used in the ABCD dataset is shown in Appendix **Table S1**). *School performance*: grades from school during last year were used as an index for school performance at both FL2 and FL3. The school performance scale is inverted so that a higher score means higher performance in school. Note that cognitive performance and mental health measures were not collected at FL3.

### Brain measures

A subset of Children (n = 7675) had 4 standardized resting-state functional MRI scans and 1 structural MRI scan at FL2. The acquired images underwent processing and quality control at the Data Analysis, Informatics, and Resource Centre of the ABCD study. Detailed protocols and processing steps can be found elsewhere (Hagler et al., 2019). To determine the resting-state functional connectivity (rs-FC) of cortical networks, the average Fisher-transformed correlation between the time courses of each pair of regions within or between 12 cortical networks defined by the Gordon atlas (Gordon et al., 2016) was calculated. Additionally, rs-FC between the 12 cortical networks and 19 subcortical regions was also computed. In total, there were 306 unique functional connectivity measures. Grey matter volume, a structural measure, was extracted from 148 regions based on the Destrieux parcellation.

### Covariates included in propensity score matching

We included the following covariates in PSM (appendix 1.1): (1) basic demographic characteristics of a participant (age in months, sex at birth, the interaction between age and sex, race, and study sites); (2) theoretically relevant factors, including parent education level and household income, pubertal status at Baseline (assessed by ABCD Youth Pubertal Development Scale and Menstrual Cycle Survey History), and BMI at baseline; and (3) confounding sleep measures, including sleep duration and total sleep disturbance score. It should be noted that we used both pubertal status/BMI at baseline as proximate for corresponding measures at FL2, due to the high percentage of missing data (at FL2, 87.1% and 26.4% of the included children have missing values on the pubertal status and BMI, respectively).

### Statistical analysis

We conducted PSM to control for covariates in the observational data, allowing us to estimate the causal effects of SJLsc on the outcome measures. The MatchIt R package (version 4.3.0) was utilized for the propensity score matching process. Participants were matched based on their probability of being in a comparison group, taking into account observed covariates using logistic regression. These above-mentioned covariates were included. The matching process involved pairing participants with low SJLsc (SJLsc <= 1) with those with high SJL (SJLsc > 1) through one-to-one matching without replacement within a predefined propensity score radius (caliper = 0.1). To assess the quality of the matching, we examined the standardized mean difference of covariates between the sufficient sleep and insufficient sleep groups and found that all covariates were well-balanced between the groups after matching (appendix 1.1), indicating that any additional group differences could not be attributed to these covariates.

For assessing these group differences an independent-sample t-test was employed, as it tends to provide a more conservative estimate of effect size compared to a paired-sample t-test (Chan et al., 2016). Subsequently, we examined how SJLsc influenced 39 behavioral domains that represented various aspects of adolescent behavior problems (e.g., aggression and rule-breaking; 20 items), cognitive functions (e.g., crystallized intelligence; six out of ten items available at FL2), mental health (e.g., psychosis and impulsivity; 12 items), and school performance at FL2.

Regarding the analysis of fMRI data, our focus was on participant pairs that had passed quality control for each corresponding brain measure. Additionally, we regressed out mean frame-wise displacement and the number of fMRI time points remaining after preprocessing from network connectivity measurements, and we also regressed out the total intracranial volume from grey matter volume measures. Multiple comparisons were corrected based on the false discovery rate (FDR) *p* < 0.05.

Based on the conceptual framework suggesting that brain measures could act as mediators in the relationship between sleep patterns and developmental outcomes, we conducted additional tests to examine whether brain measures mediate the impact of SJLsc on behavioral measures at FL2, while controlling for the covariates used in the PSM process. Our main focus was on brain and behavior assessments that exhibited a Cohen’s d greater than 0.15 between the high SJLsc and low SJLsc groups. This threshold represents a 50% increase in effect size compared to the typical range identified in the ABCD dataset (Cohen’s d 0.03–0.09) (Owens et al., 2021). By using effect size as a threshold, we aimed to enhance the replicability of neuroimaging findings.

To perform these analyses, we utilized a well-established neuroimaging mediation toolbox. Detailed processes can be found in Appendix 1.2. The significance of the mediation analyses was determined through bootstrapping with 10,000 randomly generated samples, providing a robust assessment of the results. Additional longitudinal mediation analyses were performed to test whether brain measures at FL2 mediate the relationship between SJLsc at FL2 and behavior measures at FL3 while controlling for the corresponding behavior measures at FL2. These longitudinal time-lagged analyses aim to test whether the identified brain measures can serve as biomarkers of behavioral changes over time.

Next, we replicated the above-mentioned analyses using oSJLsc. Given that only a subset of children participated in the Fitbit data collection, we applied partial correlation to examine the association between oSJLsc and behavior and brain measurements.

## Results

Of the 10414 participants included in the FL2 data, 1262 were excluded due to having a least one missing value on the covariates used in the PSM, and additional 299 were excluded for quality control for sleep duration data. After exclusion, 8853 eligible participants were included in this study, 4210 (47.5%) of whom were female, and 4643 (52.5%) were male. There were 4467 participants in the low SJLsc group and 4386 participants in the high SJLsc group. After PSM, we identified 3366 matching pairs at FL2, of which 1277 pairs had behavioral data at FL3.

We next tested how high SJLsc influenced the 43 behavioral measurements. Four cognitive measurements related to fluid intelligence were not tested in FL2. We found that high SJLsc had significant effects on 28 out of the remaining 39 available measures (FDR corrected *p* < 0.05, **see Figure 1**). Among these, seven of them had an effect size larger than 0.15, including picture vocabulary, picture sequence memory, oral reading recognition, crystallized intelligence (CI, it is calculated as a sum score of picture vocabulary and oral reading recognition, and it reflects knowledge that you have learned and stored), PPS total number of yes answers, positive urgency (PU), and grades. Surprisingly, contradictory to the previous research (Díaz-Morales & Escribano, 2015; Tamura et al., 2022), we found that high SJLsc was associated with low mood problems, i.e., depression and anxiety, with low effect size (FDR corrected *p* < 0.05, Cohen’s d < 0.1).

**Figure 1.**
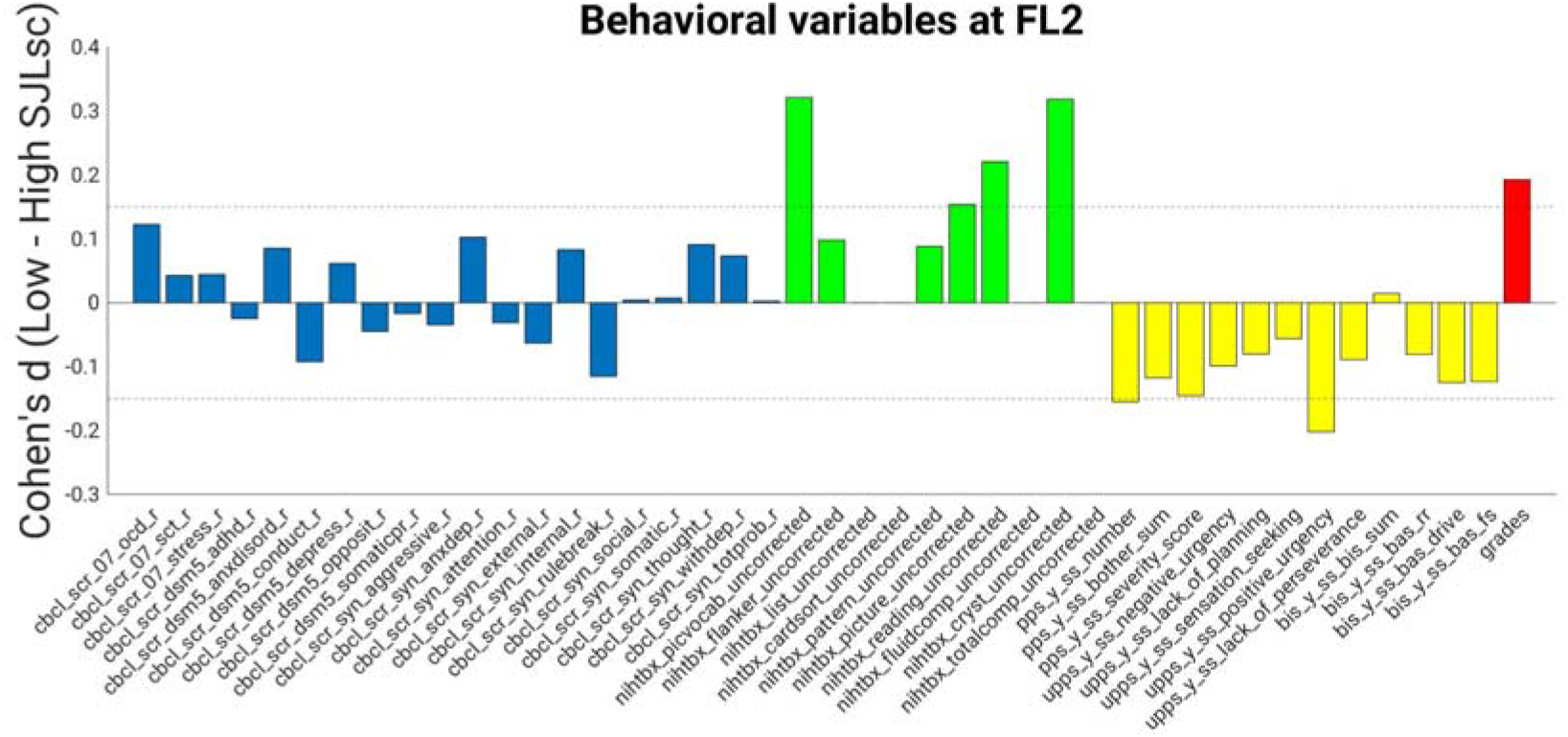
The effects of SJLsc on behavioral measures. Cohen’s d for behavior problems (blue), cognition (green), mental health (yellow), and school performance (red) in the comparison between high SJLsc and Low SJLsc groups at FL2. Positive Cohens’d value means the low SJLsc group has a higher value than that of the high SJLsc group. Note: At FL2, four measures of cognition were not available: nihtbx_list_uncorrected, nihtbx_cardsort_uncorrected, nihtbx_fluidcomp_uncorrected, and nihtbx_totalcomp_uncorrected.

At FL3, only 21 out of 43 measures were obtained. Among these, seven measures were significantly affected by SJLsc (FDR corrected *p* < 0.05). Of note, high SJLsc at FL2 is associated with low grades at FL3 (FDR corrected *p* < 0.05, Cohen’s d = 0.19 See **Figure S2**).

Out of 3366 pairs, 1864 pairs had fMRI data available. We next examined how SJLsc affects the intrinsic functional organization of brain networks in the developing brain at FL2. We found that 138 of 306 unique network connectivity measurements showed significant differences between high SJLsc and low SJLsc (See **Figure 2**). Among them, the top 5 connections (all have Cohen’s d value higher than 0.2) were cingulo-opercular - right hippocampus (cerc-hprh), cingulo-opercular - right amygdala (cerc-agrh), cingulo-parietal - left ventral DC (copa-vtdclh), sensorimotor hand - right caudate (smh-cderh), and sensorimotor hand - right putamen (smh-ptrh). Meanwhile, no gray matter volume measurements had Cohen’s d value higher than 0.1 (all FDR correct *p*’s > 0.05).

**Figure 2.**
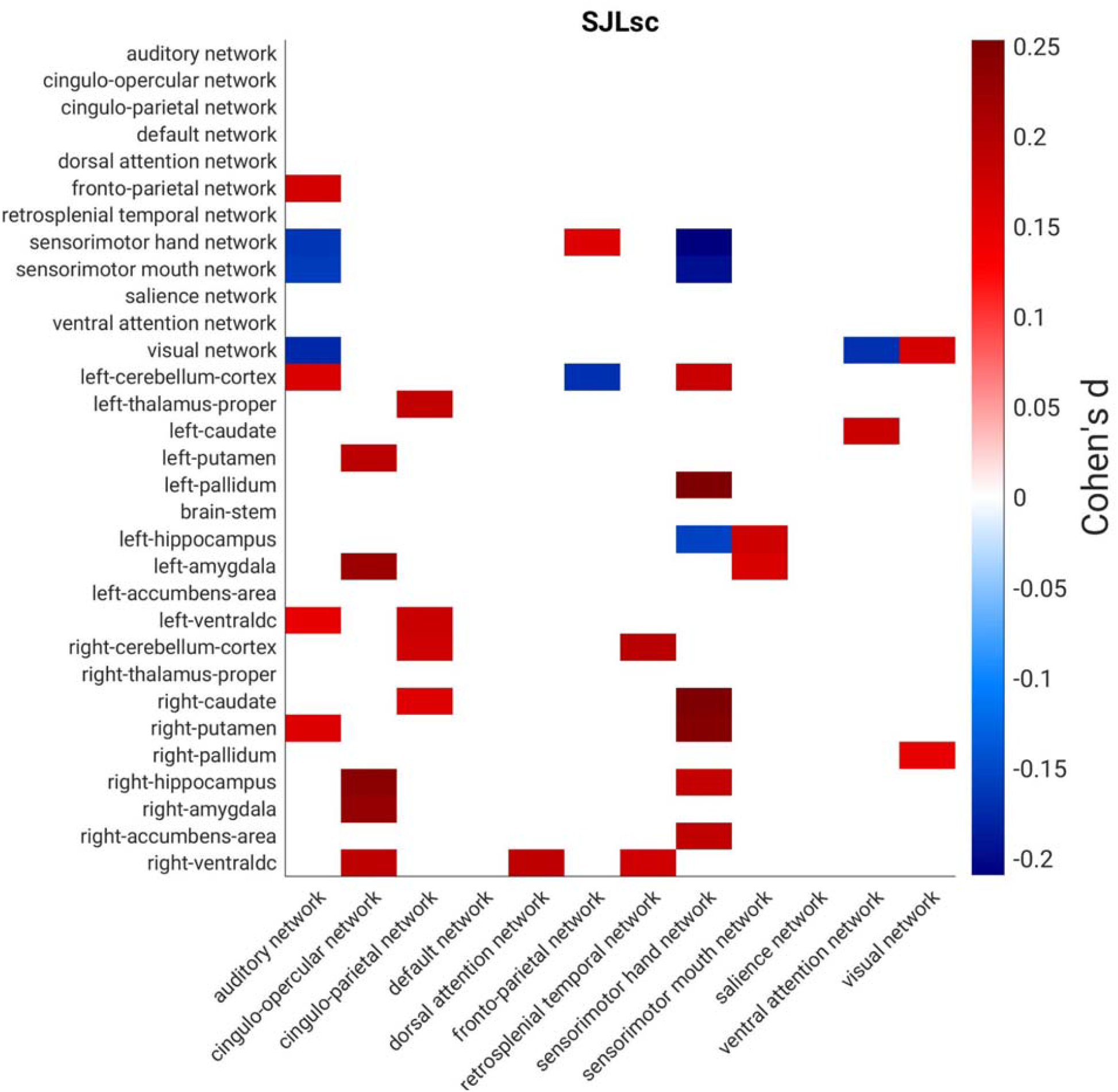
The effects of SJLsc on resting-state functional connectivity. Cohen’s d for resting-state functional connectivity measures in the comparisons between high SJLsc and low SJLsc group at FL2. Red denotes the Low SJLsc group has higher connectivity than the High SJLsc group, while blue means the High SJLsc group has lower connectivity compared to the Low SJLsc group. Note: Only connections that had Cohen’s d higher than 0.15 are shown in the figure. Within-network connectivity (i.e., visual network - visual network) is calculated by averaging unique functional connectivity pairs between all the regions in the network.

Next, to assess how network connections mediate the relationship between SJLsc and behavioral outcomes, we tested whether the five selected network connections mediated the effects of SJLsc on the seven behavioral measures identified with a Cohen’s d greater than 0.15. In this analysis, we regressed out covariances used in the PSM plus two rs-FC quality control indexes (mean motion and the number of fMRI time points remaining after preprocessing). We found that all five network connections significantly mediated the association between SJLsc and Picture-vocabulary, reading, CI, and grades (*p*’s < .05 using bootstrap sampling with 10,000 random-generated samples, see **Figure 3**). While three out of five network connections mediated the association between SJLsc and picture memory. In terms of longitudinal mediation analyses, we found that both cerc-hprh and smh-cderh at FL2 mediated the effect of SJLsc at FL2 on grades at FL3 (*p*’s < .05 using bootstrap sampling with 10,000 random-generated samples, see **Figure S3**), controlling for grades at FL2.

**Figure 3.**
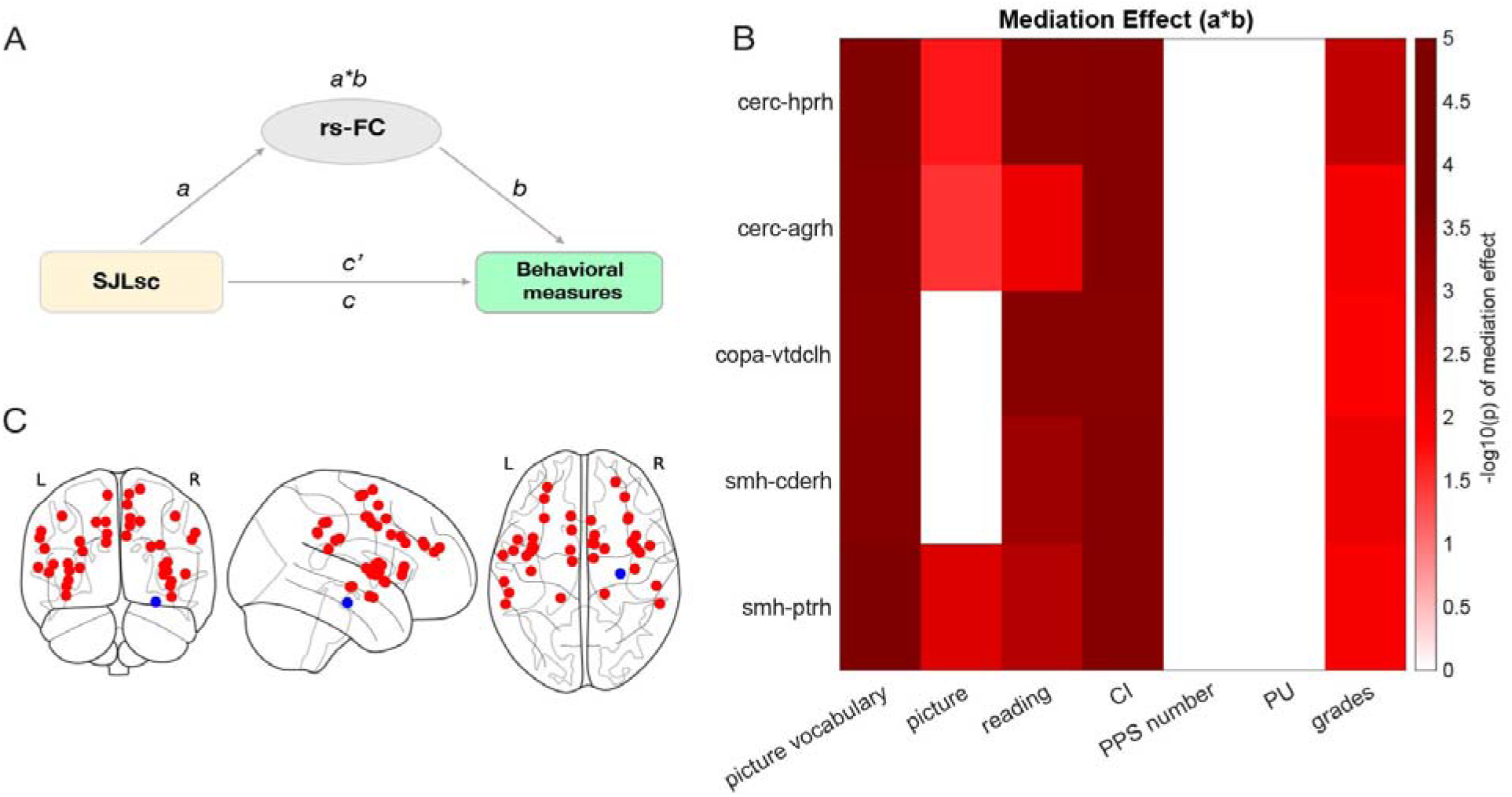
Brain measures mediated the effect of SJLsc on cognition, positive urgency, and grades. (A) Diagram of the mediation models. (B) Based on matched group comparisons, we have examined how brain measures identified with larger effect sizes (i.e., 5 rs-FC Y-axis) would mediate the effects of SJLsc on the 6 behavioral measures identified with a Cohen’s d greater 0.15 (i.e., picture vocabulary, picture memory, reading, CI, PPS number, PU, and grades; X-axis). Color bars are coded based on log-transferred bootstrapped *p*-values of the mediation effects. Only the effects reached statistical significance (*p* < .05) using bootstrap sampling with 10,000 random-generated samples shown in the figure. (C) Visualization of nodes in the cerc (red) and hprh (blue). Note: rs-FC: resting-state functional connectivity; cerc: cingulo-opercular network; copa: cingulo-parietal network; smh: sensorimotor hand network; hprh: right hippocampus; agrh: right amygdala; vtdlh: left ventral diencephalon; cderh: right caudate; ptrh: right putamen.

Last, out of 6732 matched children, 2615 had oSJLsc data available and passed quality control. In this subsample, we found that oSJLsc was weakly correlated with SJLsc (*r* = 0.22, *p* < 0.0001, **See Figure S4**). However, by using partialcorrelation analyses, the correlations patterns between oSJLsc and behavioral measures (n = 2615, see **Figure S5** and **Appendix 2.1**) and between oSJLsc and rs-FC (n = 1626, see **Figure S6** and **Appendix 2.1**) were quantitatively similar to the effects of SJLsc on behavior and rs-FC (**Figure 1** and **Figure 2**, respectively).

## Discussion

By controlling various important covariates linked to social jetlag, we assessed the impact of SJLsc on both behavior and brain. Unlike previous studies, we did not discover a meaningful correlation between SJLsc and affective functions such as depression. We found a connection between SJLsc and crystallized intelligence and school performance, which was most significantly mediated by the functional connectivity between the cingulo-opercular network and the right hippocampus. The association between SJLsc and school performance lasted for at least one year. Our results suggest that, in addition to sufficient sleep duration and quality, SJLsc may play a crucial and unique role in the neurocognitive development of early adolescents.

In contrast with previous research demonstrating a positive connection between SJL or SJLsc and mood disorders among adults (Jia et al., 2023) and adolescents (Henderson et al., 2019; Tamura et al., 2022), we found that after matching for other sleep measures and related covariates, the associations between SJLsc and depression or anxiety were negative instead of positive and with low effect size (effect size lower than 0.1). This aligns well with recent discoveries indicating that the associations between SJLsc and poor mood diminish when puberty status and other sleep issues are taken into consideration (Chen et al., 2022). Consequently, future investigations into social jetlag should include the control of other sleep metrics, such as sleep duration and quality, as well as additional sleep-related covariates.

We replicated the associations between SJLsc and cognition and school performance in a population-based, carefully controlled study. SJLsc had a large effect on crystallized intelligence with an effect size of around 0.3, which is three times larger than a typical effect size found on the ABCD dataset (Cohen’s d 0.03–0.09) (Owens et al., 2021). While these associations have been demonstrated before (Beauvalet et al., 2017; Roenneberg et al., 2019; Tamura et al., 2022), we further revealed that the neural basis of such effect involved the functional connection between the cingulo-opercular network and hippocampus (cerc-hprh). The cingulo-opercular network is known as a cognitive control network, e.g., controlling cognitive resources during episodic memory search tasks (Gratton et al., 2018; Sestieri et al., 2014). Hence, higher resting-state connectivity between cingulo-opercular network and hippocampus in Low SJLsc group might reflect better cognitive resource relocation, especially for memory-related tasks. A recent review paper (Lee et al., 2017) highlighted that hippocampal function in memory encoding is not fully developed during middle childhood (i.e., 6-12 years old). It is possible that Low SJLsc (no more than one hour) promotes hippocampal development and hence children with low SJLsc perform better in school and memory-related tasks, i.e., tasks measuring crystallized intelligence.

Despite the fact that there was only a weak correlation between subjective and objective SJLsc, quantitatively similar effects were found. It is common to have notable disparities between subjective and objective assessments of sleep measures (Aili et al., 2017). In addition, Aili et al., (2017) found objective sleep duration during weekdays was more reliable than that during weekends. It is possible that more data from actigraphy are needed, e.g., longer than 3 consecutive weeks in the current study, to reach a high agreement between subjective and objective SJLsc.

The current study offers several significant contributions. Firstly, through the utilization of PSM, the group comparison between individuals with low and high SJLsc was conducted while accounting for variables such as sleep duration, sleep quality, puberty status, BMI, socioeconomic status, and other relevant covariates. This approach effectively addressed the conflicting findings regarding the association between SJLsc and mental health, providing compelling evidence that SJLsc is indeed linked to crystallized intelligence and school performance. Secondly, we demonstrated SJLsc at FL2 predicted school performance one year later, and the connection between cingulo-opercular network and hippocampus mediated this time-lagged association. These results highlight early sleep intervention might have positive effects on school performance.

Several limitations should be acknowledged. First, due to a large amount of missing data (due to Covid-19) on puberty status and BMI, we used baseline measures as proximate values to those at FL2. Second, our sample only had limited longitudinal data on school performance. While we demonstrated a consistent association between SJLsc and grades at both FL2 and FL3, it would be valuable for future studies to explore the longitudinal association between SJLsc and other variables as more data becomes available in the ABCD study. Third, with data mostly from one time point, the current study is not able to distinguish within-person and between-person effects. A random-intercept cross-lagged panel model can mitigate this issue when more visits are available in future ABCD data releases.

The current study estimates the effects of high SJLsc (> 1h versus <=1h) on neurocognitive development in early adolescence while carefully controlling for key covariates, including other sleep measures, socioeconomic status, puberty status, and physical health indicators. Our results indicate that the connectivity between the cingulo-opercular network and the right hippocampus plays a significant role in mediating the effects of SJLsc on cognitive function and school performance. These effects are likely to last at least one year. Considering the prevalence of SJLsc and its negative consequences, early interventions such as extending sleep duration on school days (Dewald-Kaufmann et al., 2013) are necessary to enhance neurocognitive development outcomes in early adolescents.

## Data Availability

All data produced in the present study are available upon reasonable request to the authors

## Acknowledgment

We thank the ABCD consortium and NIH for providing the data for performing the research in this work. Data used in the preparation of this article were obtained from the ABCD Study (https://abcdstudy.org/) and are held in the NIMH Data Archive. This is a multisite, longitudinal study designed to recruit more than 10,000 children aged 9–10 and follow them over 10 years into early adulthood. The ABCD Study is supported by the National Institutes of Health (NIH) and additional federal partners under award numbers U01DA041022, U01DA041028, U01DA041048, U01DA041089, U01DA041106, U01DA041117, U01DA041120, U01DA041134, U01DA041148, U01DA041156, U01DA041174, U24DA041123, and U24DA041147. A full list of supporters is available at https://abcdstudy.org/federal-partners/. A listing of participating sites and a complete listing of the study investigators can be found at https://abcdstudy.org/principal-investigators/. ABCD consortium investigators designed and implemented the study and/or provided data but did not necessarily participate in the analysis or writing of this report. This manuscript reflects the views of the authors and may not reflect the opinions or views of the NIH or ABCD consortium investigators. We are not paid to write this article by a pharmaceutical company or other agency.

## Author contributions

F.N.Y. conceptualized the study, analyzed the data, generated figures, and wrote the original draft. D.P. edited the manuscript. J.H.D. supervised the study and edited the manuscript.

## Competing interests

The authors declare no competing interests.

## Ethics approval statement

ABCD study received ethical approval in accordance with the ethical standards of the 1964 Declaration of Helsinki.

## Notes

### Competing Interest Statement

The authors have declared no competing interest.

### Author Declarations

The study used ONLY openly available human data that were originally located at nda.nih.gov/abcd

